# Antibody prevalence after 3 or more COVID-19 vaccine doses in 23,000 immunosuppressed individuals: a cross-sectional study from MELODY

**DOI:** 10.1101/2023.02.09.23285649

**Authors:** Fiona A Pearce, Sean H Lim, Mary Bythell, Peter Lanyon, Rachel Hogg, Adam Taylor, Gillian Powter, Graham S Cooke, Helen Ward, Joseph Chilcot, Helen Thomas, Lisa Mumford, Stephen P McAdoo, Gavin J Pettigrew, Liz Lightstone, Michelle Willicombe

## Abstract

**Objectives:** To investigate the prevalence of spike-protein antibodies following at least 3 COVID-19 vaccine doses in immunocompromised individuals.

**Design:** Cross-sectional study using UK national disease registries of individuals with solid organ transplants (SOT), rare autoimmune rheumatic diseases (RAIRD) and lymphoid malignancies (LM).

**Setting:** Participants were identified, invited and recruited at home by accessing the NHS Blood and Transplant Registry for those UK individuals who had received a SOT; and the National Disease Registration Service at NHS Digital for identifying individuals within England with RAIRD or LM.

**Participants:** 101972 people were invited, 28411 recruited, and 23036 provided serological data, comprising 9927 SOT recipients, 6516 with RAIRD, and 6593 with LM.

**Interventions:** Participants received a lateral flow immunoassay for spike-protein antibodies to perform at home together with an online questionnaire.

**Main outcome measures:** Odds of detectable IgG spike-protein antibodies in immunosuppressed cohorts following at least three COVID-19 vaccine doses by participant demographic, disease type, and treatment related characteristics

**Results:** IgG spike-protein antibodies were undetectable in 23.3%, 14.1% and 20.7% of the SOT, RAIRD and LM cohorts, respectively. Participants had received three, four or ≥five vaccine doses at the time of testing in 28.5%, 61.8%, and 9.6%, respectively. In all groups, seropositivity was associated with younger age, higher number of vaccine doses and previous COVID-19 infection. Immunosuppressive medication reduced the likelihood of seropositivity: the lowest odds of seropositivity were found in SOT recipients receiving an anti-proliferative agent, calcineurin inhibitor and steroid concurrently, and those treated with anti-CD20 in the RAIRD and LM cohorts.

**Conclusions:** Approximately one in five individuals with SOT, RAIRD and LM have no detectable IgG spike-protein antibodies despite three or more vaccines, but this proportion reduces with sequential booster doses. Choice of immunosuppressant and disease-type is strongly associated with serological response. Antibody testing could enable rapid identification of individuals who are most likely to benefit from additional COVID-19 interventions.

**Trial registration:** Clinicaltrials.gov, NCT05148806

## Introduction

Despite vaccination, immunocompromised people have poorer outcomes from COVID-19 infection than observed in the general population^1-6^. Population-level analyses have highlighted that those with solid organ transplants, lymphoid malignancies such as lymphoma, and individuals on immunosuppressive medication have the highest risk of hospitalisation or death from COVID-19^2,4^. These groups were included in the WHO definition of moderately to severe immunocompromised persons, in whom, an extended primary vaccine series was recommended^7^. Aligned with WHO recommendations, these populations have since been prioritised for additional booster vaccinations, and in the UK, for COVID-19 therapeutics in the community^8^. However, the groups defined are broad, and at an individual level, there is likely considerable variation in the risks of COVID-19, with vaccine immunogenicity studies to date not powered to either examine this heterogeneity or relate to disease protection^9-11^.

As it is recognised that a substantial proportion of immunocompromised people fail to mount serological responses despite multiple inoculations of SARS-CoV-2 vaccines, assessment of serological responses in this population may enable risk-stratification and facilitate personalised targeting of additional anti-viral interventions^9-13^. However, mass antibody testing has not been implemented at population level in the UK or internationally. In part this is due to the development of multiple assays with different sensitivity, but also because of the lack of definition of a correlate of protection and hence clinical application^14,15^.

The availability of a rapid, point-of-care test whose result correlates with the risk of severe infection or death within the broadly defined high risk groups would redefine our approach in managing these individuals. In response to this need, we sought to evaluate whether mass antibody testing using a lateral flow immunoassay (LFIA), when coupled with self-reporting of participant factors likely to impact immune responsiveness, could establish particular risk factors for absent antibody responses following three or more COVID-19 vaccines across three different groups of immunocompromised individuals.

## Methods

### Study design and data sources

We conducted a prospective, observational cohort study of three immunosuppressed populations: SOT recipients, people with RAIRD, and people with LM, and herein present the baseline cross-sectional data. The study design was adapted from the REACT2 study, applied to an immunocompromised population^16^. Participants were identified and invited by accessing the comprehensive UK national disease registers: NHS Blood and Transplant (NHSBT) Transplant Registry for those UK individuals who had received a SOT; and the National Disease Registration Service (NDRS) at NHS Digital for identifying individuals within England with RAIRD or LM.

### Participant recruitment and consent

The study was open to recruitment between Dec 7, 2021 to June 26, 2022. All participants had to be ≥18 years old and have had at least three COVID-19 vaccine doses at the time of recruitment. Initial recruitment of SOT recipients differed from the RAIRD and LM cohorts; this was a pragmatic decision based on differences in patient identifiable data recorded by NHSBT and NDRS registries. Self-registration of UK SOT recipients directly via the study web-portal without invitation commenced from Dec 7, 2021, with eligibility confirmed by verification of the provided NHS number (or CHI number in Scotland) against the NHSBT Transplant Registry.

From Feb 14, 2022, invitations were sent to all residents in England who had a recent diagnosis of lymphoma or multiple myeloma or who had recent hospital care for a probable diagnosis of a rare autoimmune rheumatic disease (small vessel vasculitis (SVV), systemic lupus erythematosus (SLE), myositis, systemic sclerosis or giant cell arteritis). Patients with RAIRD were identified by the NDRS data using algorithms applied to Hospital Episode Statistics, *Supplementary Data*^17^. People with lymphoid malignancies were registered in the National Cancer Registration Dataset in 2019 or in the Rapid Cancer Registration Dataset in 2020 or 2021, *Supplementary Data*^17,18^. From March 31, 2022, invitations were sent to all eligible SOT recipients in England and Wales on the UK Transplant Registry who had not previously registered.

Following invitation, participants registered via a web portal developed by Ipsos-MORI. Consent was gained electronically for participation in the study, subsequent data linkage, and willingness to be contacted about any future interventional research. At the time of registration participants were asked to complete a questionnaire including information on socio-demographic variables, vaccination and COVID-19 infection history, clinical diagnoses and immunosuppression treatment. Following registration into the study, participants were sent a LFIA test with instructions, and asked to read and report the result on the study web portal, uploading a photograph where possible. Participants then completed a second short online questionnaire on shielding history, psychological distress, experience of the test and the test result. Self-reported depression and anxiety was evaluated using the PHQ-8 and GAD-7 scores, which were combined to form a composite measure of psychological distress (PHQ-ADS)^19^.

### Rapid antibody testing for SARS-CoV-2

The Fortress COVID-19 Total Antibody LFIA device was used for the detection of IgM and IgG antibody directed against the SARS-CoV-2 Spike (S) protein; it has a reported sensitivity of 92% and specificity of 95% in solid organ transplant recipients^16^. Participants who tested positive for IgG only or IgG and IgM were classified as antibody positive, whereas those who positive for IgM only or tested negative were classified as antibody negative.

### Sample Size

The study planned to recruit 36000 participants (12000 in each patient population) and of these it was expected 85% (n=30600) would return a valid LFIA result. Following a third vaccine, we estimated approximately one third of patients in each group would have no SARS-CoV-2 antibodies, so the sample size would give a 95% confidence interval for the proportion of patients with no antibodies of ±0·91% for each patient group^13^. Time constraints restricted recruitment to 28411 participants, with 23036 returning a valid LFIA result for inclusion in the final cohort.

### Statistical analysis

Demographic characteristics including those common across all populations (number of vaccines at test, vaccine type, gender, ethnicity, anxiety, depression, previous COVID-19 infection, age, height, weight, body mass index (BMI), and comorbidities) and population specific were summarised, stratified by antibody status. These were for SOT: transplant type, graft number, time from transplant, cancer, rejection (12 months prior to first vaccine or from first vaccine to time of study registration), immunosuppression. For RAIRD: diagnosis, time from diagnosis, disease activity, time from last disease flare, immunosuppression, time from vaccine; and for LM: diagnosis, time from diagnosis, immunosuppression, time from vaccine. Differences in characteristics by antibody status were tested univariately using the chi-squared test for categorical variables and Kruskal-Wallis test for continuous variables.

Multivariable analysis was undertaken using a binary logistic regression model to identify which factors were independently associated with developing antibodies. Regression models were performed for each cohort individually. We conducted a complete case analysis, excluding individuals with missing variables, except for those with an unknown previous COVID-19 infection, as we could not assume this was missing at random. A stepwise variable selection process was used to identify factors, using a 10% significance level for inclusion and to subsequently remain in the model, and individual variables in the model should be interpreted in the presence of all other factors in the model. A sub-analysis was undertaken by adding psychological distress to the multivariable model to identify if it was independently associated with developing antibodies. This was not part of the main analysis due to a large proportion of missing data, which was also expected to be missing not at random. To assess the ability of the model to discriminate patients who had a positive antibody status, concordance statistics (C-statistic; equal to the area under the receiver operating curve) and 95% CIs were calculated. Assessment of the standardised Pearson residuals were used for model checking.

### Ethics and legal basis

Participants with LM and RAIRD were identified within the NDRS, under the National Disease Registries Directions 2021, made in accordance with sections 254(1) and 254(6) of the 2012 Health and Social Care Act. Participant with SOT were identified by NHSBT under Paragraph 7 of Chapter 2 to the Data Protection Act 2018 which says that, as a government body, NHSBT may process personal data as necessary for the effective performance of a task carried out in the public interest. Ethical approval for MELODY (Mass Evaluation of Lateral flow immunOassays for the Detection of SARS-CoV-2 antibodY responses in immunosuppressed people) was granted by London Central Research Ethics Committee (21/HRA/4858) on 21 November 2021. Confidentiality Advisory Group (‘section 251 support’) to process confidential patient information without consent for the purpose of sending personal invitation letters (22/CAG/0004) was granted on 10 December 2021. National data opt outs, and NDRS opt outs were applied. This study is registered with clinicaltrials.gov, NCT05148806

### Role of the funding source

The study is funded by the MRC (MR/W029200/1), Kidney Research UK, Blood Cancer UK, Vasculitis UK and the Cystic Fibrosis Trust. The funders of the study had no role in study design, data collection, data analysis, data interpretation, or writing of the report.

## Results

A total of 98725 individuals in the NHSBT and NDRS Registries were invited to participate, and 3247 SOT recipients self-enrolled into the study, giving recruitment of 28411 participants, of whom 23036 returned a valid LFIA result (9927 participants with a SOT, 6516 with a RAIRD and 6593 with a LM; Figure 1). The response rate to invitations was 25.5%, and details of how the recruited cohort compares to the whole populations made be found in the *Supplementary Data*.

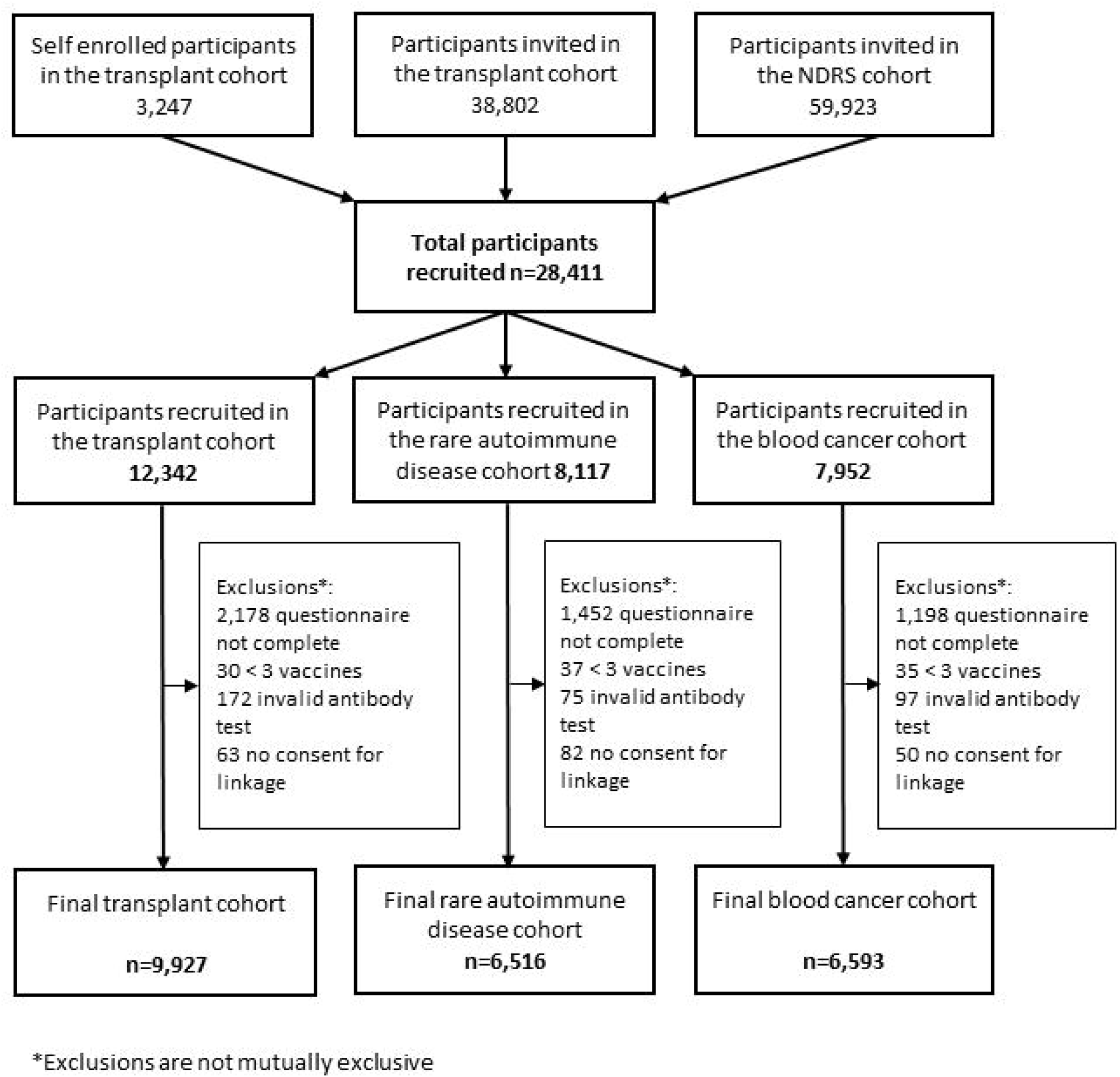

The demographic and vaccine series data of the SOT, RAIRD and LM cohorts are detailed in Table 1. Participants had received three, four or ≥five vaccine doses at the time of testing in 6583 (28·6%), 14234 (61·8%), and 2219 (9·6%), respectively; and confirmed prior infection was reported in 3113 (31·4%), 1920 (29·5%) and 1692 (25·7%) of SOT, RAIRD, and LM participants, respectively. Home LFIA testing identified a positive antibody response in 7617 (76·7%) SOT recipients, compared to 5594 (85·8%) in the RAIRD, and 5227 (79·3%) in the LM cohorts, Table 1.

**Table 1.**
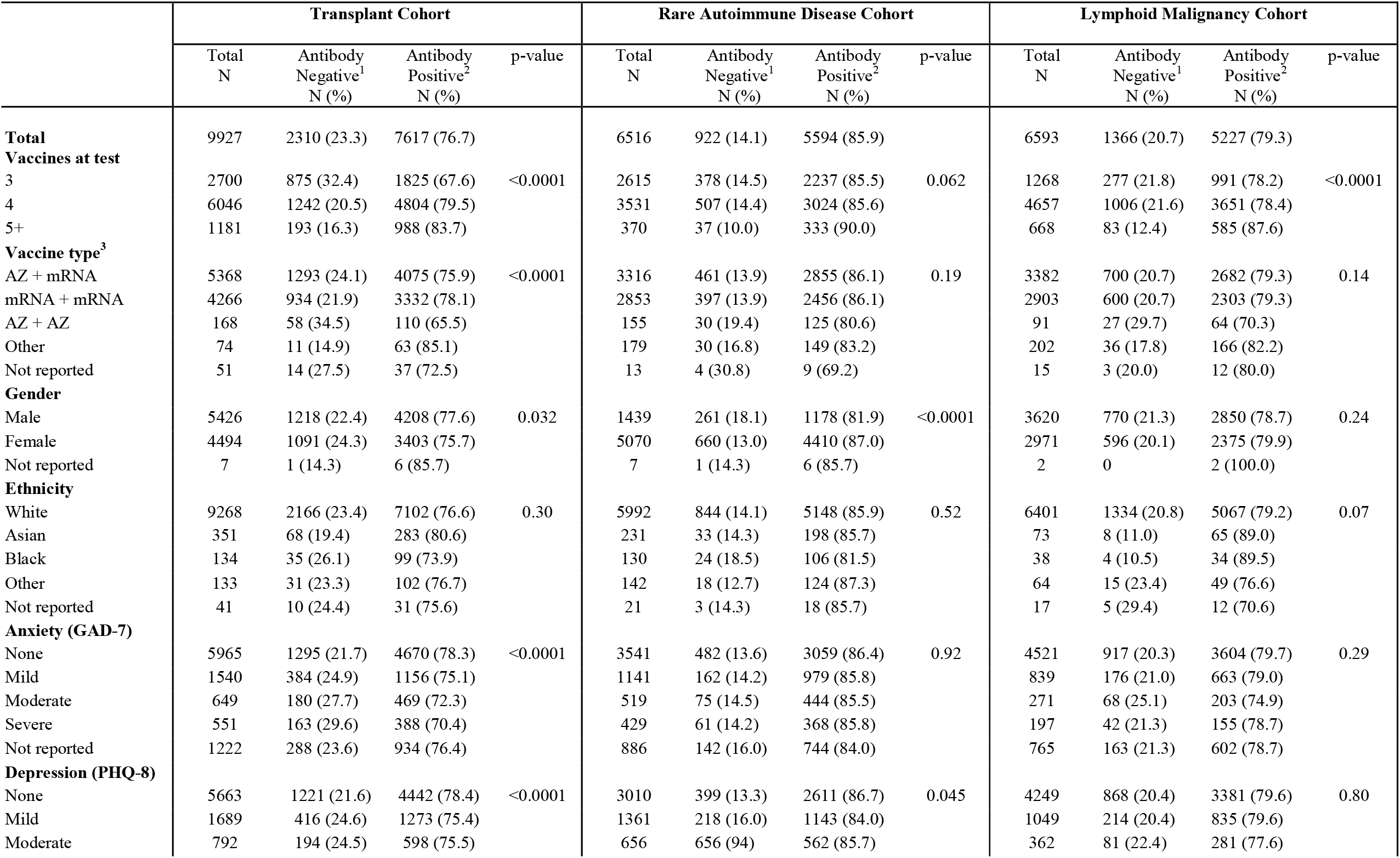
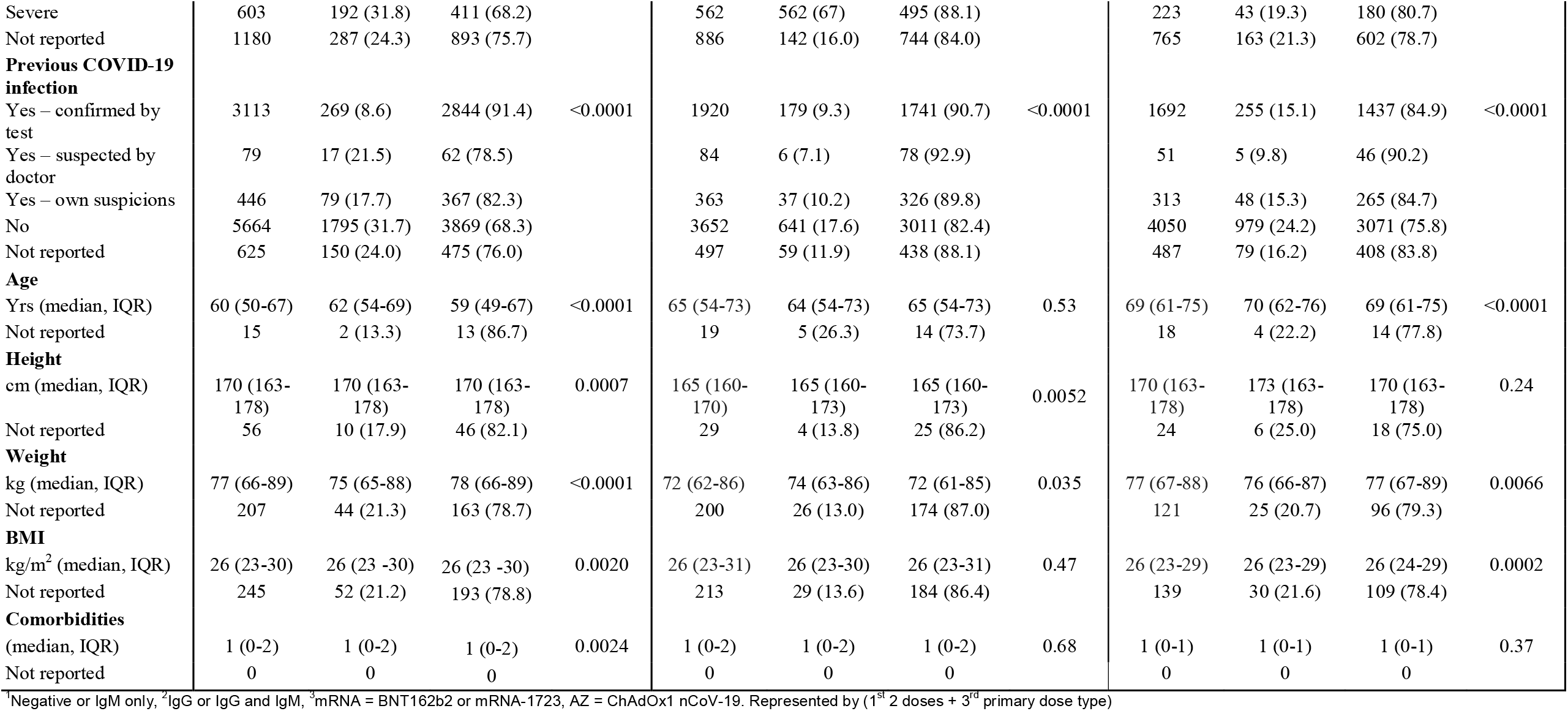
Demographic characteristics by antibody status.

The majority of participants within the SOT cohort had received one prior transplant: 6591 (66·5%) kidney, 1981 (20·0%) liver, 596 (6·0%) cardiac, and 9419 (94·9%) were ≥1 year post-transplant at receipt of latest vaccine dose, Table 2. Regarding immunosuppression, 6121 (61·7%) were prescribed both an antiproliferative agent and a calcineurin inhibitor (CNI), and 4876 (49·1%) were receiving steroids as part of their treatment regimen. Rejection episodes had occurred in 199 (2·0%) participants.

**Table 2.** Demographic characteristics of solid organ transplant recipients who had 3 or more vaccines by antibody status.

|  | **Total** | **Antibody**<br>**negative<sup>1</sup>**<br>**N (%)** | **Antibody**<br>**positive<sup>2</sup>**<br>**N (%)** | **p-**<br>**value** |
| --- | --- | --- | --- | --- |
| **Total** | 9927 | 2310 (23.3) | 7617 (76.7) |  |
| **Transplant Type** |  |  |  |  |
| Kidney only | 6591 | 1601 (24.3) | 4990 (75.7) | <0.0001 |
| Liver only | 1981 | 327 (16.5) | 1654 (84.5) |  |
| SPK/ Panc/ Islet/ SIK | 350 | 88 (25.1) | 262 (74.9) |  |
| Heart only | 596 | 153 (25.7) | 443 (74.3) |  |
| Lung (including heart-lung) | 333 | 118 (35.4) | 215 (64.6) |  |
| Other | 76 | 23 (30.3) | 53 (69.7) |  |
| **Graft number** |  |  |  |  |
| First graft | 8696 | 1991 (22.9) | 6705 (77.1) | 0.021 |
| Re-graft | 1231 | 319 (25.9) | 912 (74.1) |  |
| **Cancer diagnosis since transplant** |  |  |  |  |
| No | 8599 | 2001 (23.3) | 6598 (76.7) | 0.74 |
| Yes | 1276 | 291 (22.8) | 985 (77.2) |  |
| Not reported | 52 | 18 (34.6) | 34 (65.4) |  |
| **Rejection** |  |  |  |  |
| No | 9648 | 2220 (23.0) | 7428 (77.0) | <0.0001 |
| Yes - prior to vaccine | 88 | 17 (19.3) | 71 (80.7) |  |
| Yes - post vaccine | 86 | 39 (45.3) | 47 (54.7) |  |
| Yes – prior and post vaccine | 25 | 9 (36.0) | 16 (64.0) |  |
| Not reported | 80 | 25 (31.3) | 55 (68.8) |  |
| **Immunosuppression** |  |  |  |  |
| Belatacept based only | 1 | 0 | 1 (100.0) | <0.0001 |
| Antiproliferative and Calcineurin only | 3274 | 740 (22.6) | 2534 (77.4) |  |
| Antiproliferative only | 199 | 32 (16.1) | 167 (83.9) |  |
| Calcineurin Inhibitor only | 1467 | 191 (13.0) | 1276 (87.0) |  |
| Other only | 92 | 9 (9.8) | 83 (90.2) |  |
| Belatacept based and Steroid | 10 | 5 (50.0) | 5 (50.0) |  |
| Antiproliferative and Calcineurin and Steroid | 2847 | 891 (31.3) | 1956 (68.7) |  |
| Antiproliferative and Steroid only | 434 | 98 (22.6) | 336 (77.4) |  |
| Calcineurin Inhibitor and Steroid only | 1434 | 318 (22.2) | 1116 (77.8) |  |
| Other and Steroid | 151 | 25 (16.6) | 126 (83.4) |  |
| None | 18 | 1 (5.6) | 17 (94.4) |  |
| Not reported |  | 0 | 0 |  |
| **Time from transplant to most recent vaccine** |  |  |  |  |
| Vaccine pre-transplant | 21 | 6 (28.6) | 15 (71.4) | 0.84 |
| 0-89 days post-transplant | 52 | 14 (26.9) | 38 (73.1) |  |
| 90-364 days post-transplant | 435 | 104 (23.9) | 331 (76.1) |  |
| ≥1 year post-transplant | 9419 | 2186 (23.2) | 7233 (76.8) |  |
| Not reported | 0 | 0 | 0 |  |
| **Days from most recent vaccine to test (median, IQR)** | 89 (57-127) | 88 (60-126) | 89 (56-127) | 0.83 |
| Not reported | 0 | 0 | 0 |  |

<sup>1</sup>Negative or IgM only, <sup>2</sup>IgG or IgG and IgM; SPK – simultaneous pancreas and kidney; SIK – simultaneous islet and kidney

Of the SOT cohort, 9233 (93·0%) participants were included in the logistic regression analysis, Figure 2 and *Supplementary Data*. Older age (OR 0·70 [95% CI 0·66-0·73]) correlated with a negative antibody response, whilst number of vaccine doses received (5 versus 3, OR 2·76 [2·28-3·35]) and prior COVID-19 infection (OR 4·16 [3·65-4·75]) correlated with a positive response. Liver transplant recipients were most likely to develop antibodies (OR 1·27 [1·09-1·49]), and lung transplant recipients least likely (OR 0·59 [0·46-0·77]) compared to kidney transplant recipients. The number of prescribed immunosuppressants (rather than the particular agents) correlated with antibody response: compared to patients receiving ‘dual’ antiproliferative plus CNI therapy, the odds of detectable antibody were lower in patients on ‘triple immunosuppression’ (OR 0·61 [0·53-0·70]), but higher in those receiving either antiproliferative or CNI monotherapy (ORs 1.71 [1.11-2.64] and 2·02 [1·66-2·45] respectively). A documented rejection episode and receipt of a ChAdOx1 nCoV-19 vaccine was also associated with absent antibody responses (ORs 0·51 [0·36-0·71] and 0·86 [0·78-0·96] respectively).

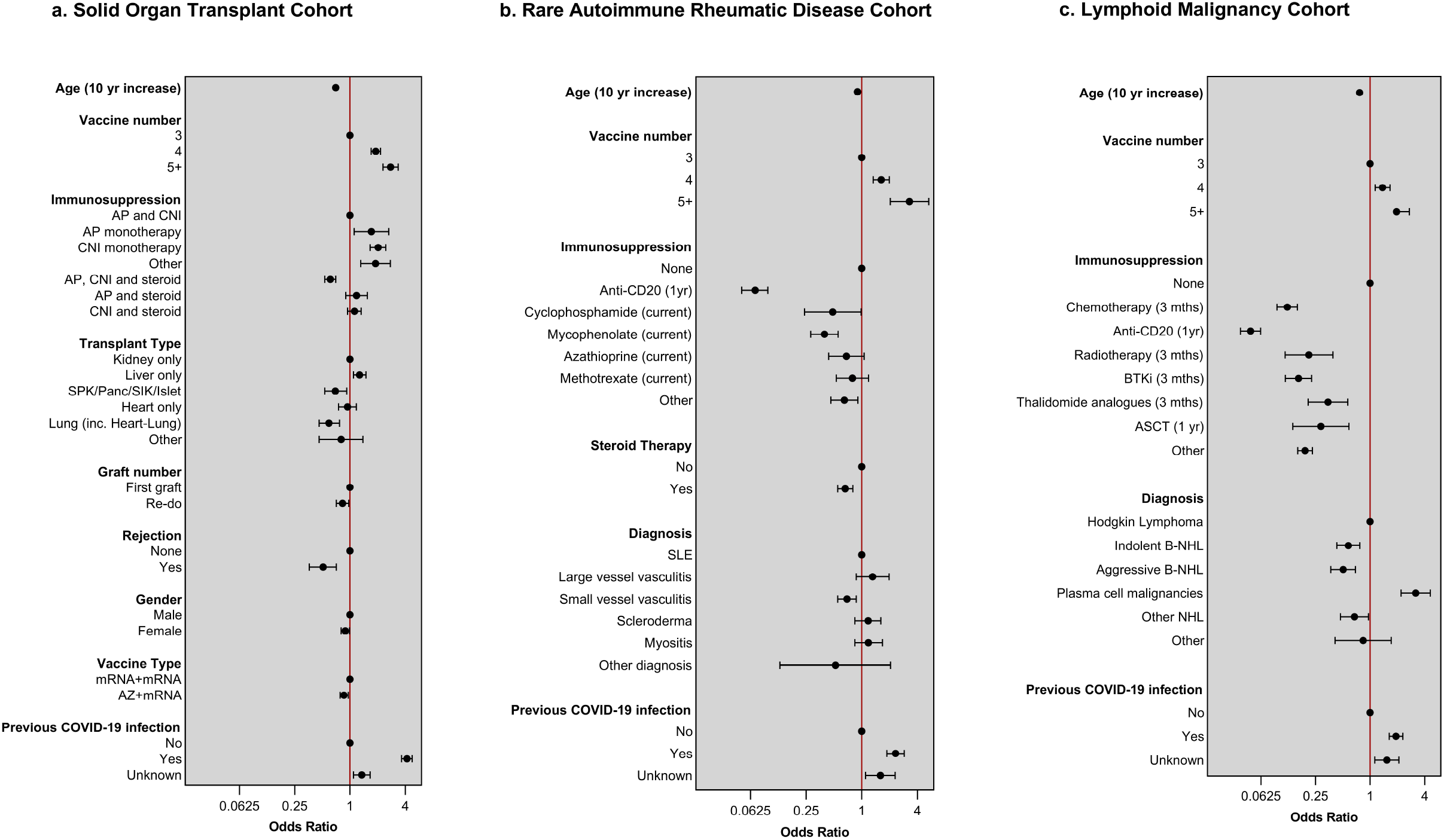

The RAIRD cohort comprised participants with SLE (2412, 37·0%), SVV (1364, 20·9%), scleroderma (869, 13·3%) and large vessel vasculitis (574, 8·8%). The majority (3263, 59·3%) had longstanding disease, diagnosed ≥5 years prior to recruitment, Table 3. Most participants (4136, 63·5%) reported mild or moderate disease activity, whereas 287 (4·4%) reported severe disease activity at recruitment, and 2160 (33·1%) had experienced a disease flare ≤1 year prior to, or following, first vaccination. The majority (4231, 64·9%) were currently receiving immunosuppressive medication and 2479 (41·7%) were receiving steroids, Table 3.

**Table 3.**
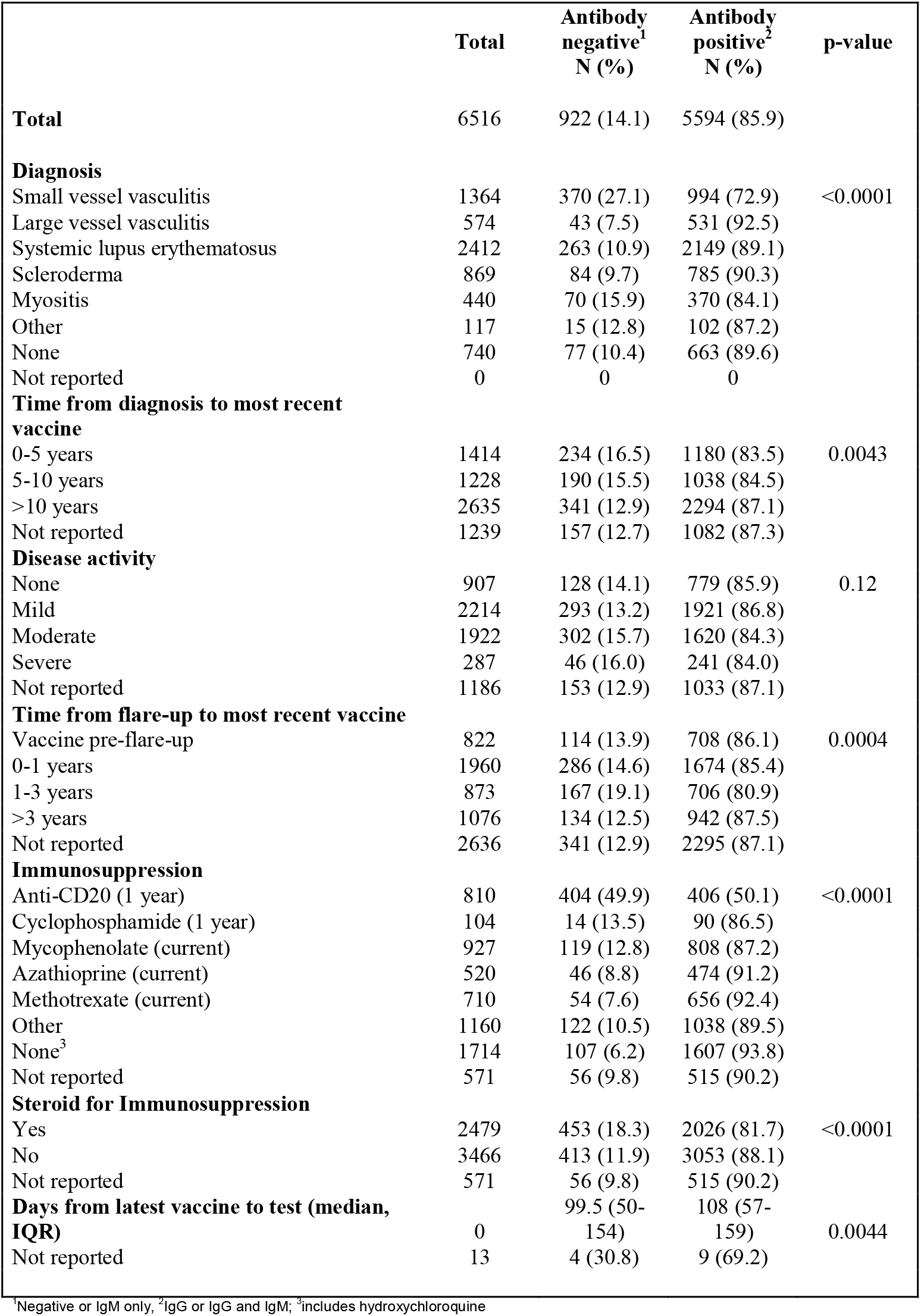
Demographic characteristics of participants with rare autoimmune disease who had 3 or more vaccines by antibody status.

Of the RAIRD cohort, 4866 (74·7%) participants were included in the logistic regression analysis, Figure 2 and *Supplementary Data*. After adjustment, older age correlated with a negative antibody response (OR 0·90 (0·83-0·97]), whereas increasing number of vaccine doses (5 versus 3, OR 3·28 [2·03-5·30]) and prior COVID-19 infection (OR 1·59 [1·10-2·30]) correlated with a positive antibody response, in keeping with findings in the SOT cohort. Amongst disease subtypes, participants with SVV were least likely to have a positive antibody response (OR 0·69 [0·55-0·87]) independent of all other factors. Current or recent receipt of any immunosuppression was associated with a reduced odds of positive antibody response compared to no immunosuppression (which included hydroxycholoroquine-only), but to a variable extent: anti-CD20 within the last year had greatest impact with a >90% reduced odds of a positive antibody response (OR 0·07 [0·05-0·10]); cyclophosphamide in the last year and current mycophenolate conferred a 50-60% reduced odds (ORs 0·49 [0·24–0·98] and 0·39 [0·28–0·55] respectively); whereas current azathioprine, methotrexate or other biologic/conventional immunosuppressants had modest effects (estimated at a 20-30% reduced odds of positive antibody response but all being of borderline statistical significance). Current steroid treatment was also associated with reduced odds of positive antibody response (OR 0·66 [0·55-0·80]), irrespective of other immunosuppression.

The LM cohort included participants with indolent B-cell non-Hodgkin lymphoma (B-NHL, 2706, 41·0%), plasma cell malignancies (1327, 20·1%), and aggressive B-NHL (1017, 15·4%), Table 4. Most participants (5438, 82·5%) were diagnosed within three years of latest vaccine dose. Active anti-cancer treatment was received by 2912 (44·2%) of participants, with no treatment in 3288 (49·9%). Chemotherapy (537, 8·1%), anti-CD20 (387, 5·9%), thalidomide analogues such as thalidomide, lenalidomide and pomalidomide (392, 5·9%), Bruton tyrosine kinase inhibitors (BTKi, 190, 2·9%), autologous stem cell transplantation (ASCT, 100, 1·5%) and radiotherapy (53, 0·8%) comprise the specified treatment groups.

**Table 4.**
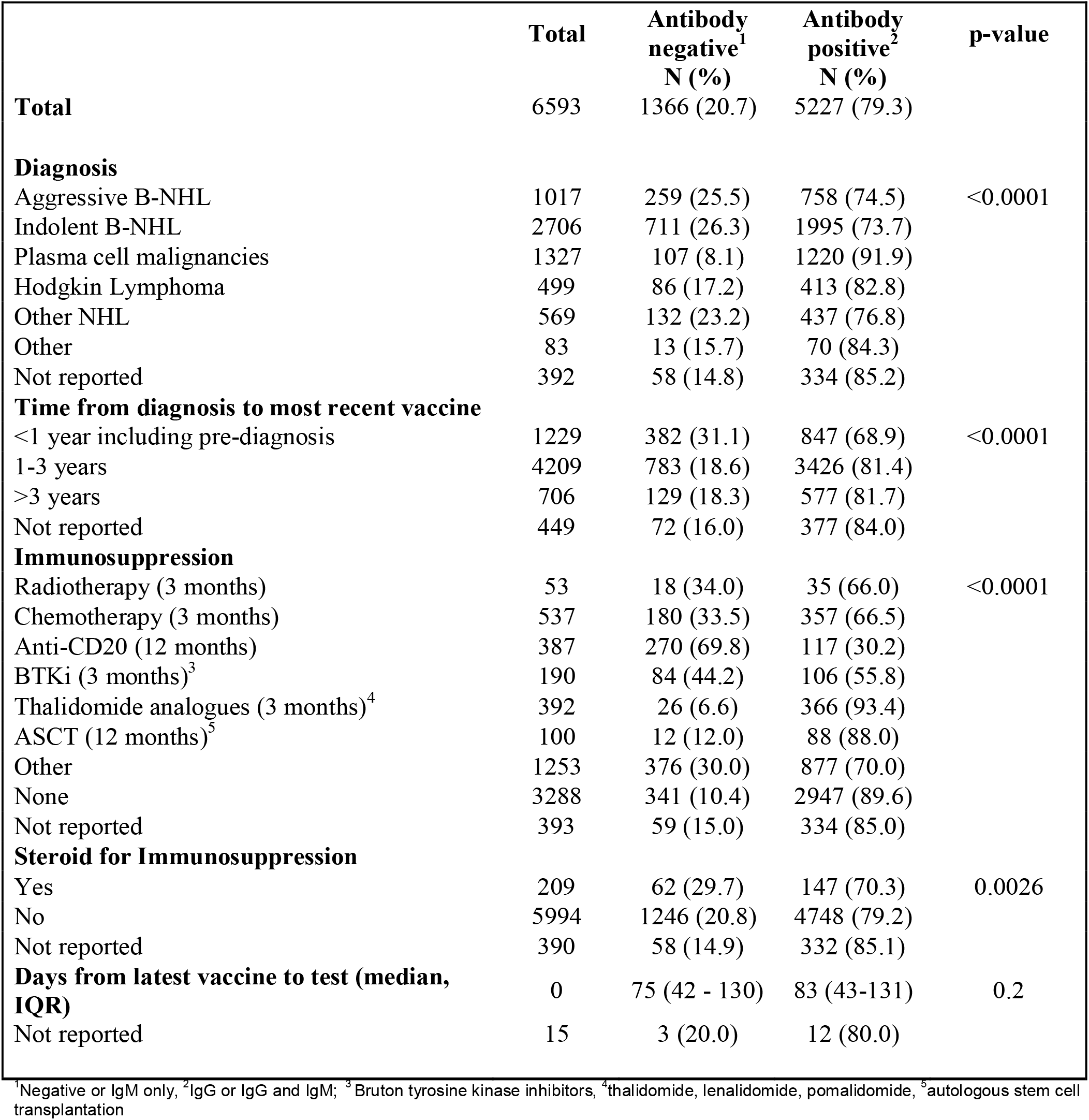
Demographic characteristics of participants with lymphoid malignancy who had 3 or more vaccines by antibody status.

Of the LM cohort, 5737 (87·0%) participants were included in the logistic regression analysis, Figure 2 and *Supplementary Data*. Like the SOT and RAIRD cohorts, older age (OR 0·76 [0·71-0·82]) correlated with a negative antibody response, whilst prior COVID-19 infection (OR 1·93 [1·63-2·29]), and vaccine dose (5 versus 3 OR 1·96 [1·42-2·71]) correlated with a positive response. Both aggressive and indolent B-NHL participants were less likely to have a positive response compared to those with Hodgkin lymphoma (ORs 0·50 [0·37-0·69], and 0·58 [0·43-0·77], respectively). In contrast, participants with plasma cell malignancies had an increased likelihood of a positive antibody response compared to Hodgkin lymphoma (OR 3·19 [2·20-4·62]). Amongst the anti-cancer treatments, anti-CD20-treated participants were least likely to develop a positive antibody response (OR 0·05 [0·04-0·06]), followed by chemotherapy (OR 0·12 [0·09-0·16]) and BTKi (OR 0·16 [0·12-0·23]), radiotherapy (OR 0·21 [0·12-0·39]), ASCT (OR 0·29 [0·14-0·58]), and thalidomide analogues (OR 0·34 [0·21-0·57]).

The influence of physical and mental health comorbidities on antibody response was also assessed in each cohort, *Supplementary Data*. Moderate-severe psychological distress was present in 12·4%, 16·9% and 7·4%, of the SOT, RAIRD and LM cohorts who completed the PHQ-ADS, respectively. In the SOT cohort, those reporting psychological distress had significantly reduced odds of a positive antibody response compared with no distress after adjustment for other significant variables (OR 0·64 [0·54-0·75]), *Supplementary Data*. Non-response to the PHQ-ADS was also associated with reduced odds of a positive antibody response (OR 0·85 [0·74-0·98]). A separate unadjusted analysis of a self-reported history of depression was associated with negative antibody responses in the SOT cohort, but no associations with other mental health conditions were observed *Supplementary Data*. Conversely, in the RAIRD cohort, psychological distress was not associated with antibody status, yet similarly to the SOT cohort, participants who did not respond to the PHQ-ADS were more likely to have negative antibody responses compared with the group reporting no distress. In the LM group, no association was found between distress and antibody status.

## Discussion

MELODY has identified the demographic, clinical and therapeutic characteristics associated with lack of detectable SARS-CoV-2 antibodies following three or more doses of COVID-19 vaccine in more than twenty-three thousand people across three distinct immunosuppressed populations. We show that it is possible to find, identify, invite and test whole cohorts of people who are immunosuppressed. Key findings are that higher number of vaccine doses, previous COVID-19 infection, and younger age are associated with increased odds of detectable SARS-CoV-2 IgG antibody in all cohorts. Our large study size has enabled estimation of the odds of detectable SARS-CoV-2 IgG antibody in more detailed subcohorts of the immunocompromised than reported previously, thereby enabling stratification by disease and treatment type. Risk stratification has three main implications: groups that have a higher risk of absent SARS-CoV-2 IgG antibody could be offered antibody testing; those with undetectable serological responses could be offered specific interventions e.g. further vaccines or pre-exposure prophylaxis; and future immunotherapies recommended for each underlying disease could be modified in the light of different risks of impaired vaccine-induced immunity. The data also reinforces the need for ongoing personal protective measures and need for all close contacts of immunocompromised persons to receive vaccines as scheduled.

Our results are consistent with prior sub-group specific immunogenicity studies that incorporated more sensitive serological assays, thereby supporting the rationale of our approach^9-12^. However, in addition, we report shared characteristics associated with antibody status across all three cohorts. As the public develops increasing vaccine fatigue, our demonstration that increased vaccine doses associates with higher rates of seropositivity in the immunocompromised will be crucial for promoting future vaccine boosting. It also presents options for developing bespoke booster vaccination schedules for this population. In addition to shared characteristics, we were also able to report on differences between cohorts, and found reduced responses following vaccine combinations that included adenovector viral vaccines in the SOT cohort only. We hypothesise that this could reflect the dominance of T-cell directed immunosuppression therapy in SOT patients, which, by impairing the robust cellular responses elicited by adenovector viral vaccines, may block critical T cell help for antibody production^20,21^.

Cohort-specific observations consistent with previous studies include the finding that in SOT recipients, immunosuppression burden is an important factor determining odds of seroconversion, with patients receiving triple immunosuppression at highest risk of remaining seronegative following vaccination^10,13,22,23^. In RAIRD, it is also accepted that patients receiving anti-CD20 therapy are less likely to have detectable SARS-CoV-2 IgG antibody post-vaccination^9,12^. Whilst in haematological malignancies, systemic anti-cancer therapy is recognised to be the biggest predictor of the humoral response to vaccination^11^. In particular, treatments such as anti-CD20 and BTKi have been observed to be strongly associated with negative seroprevalence^11^. However, all these observations have been largely derived from relatively small, heterogeneous datasets. The MELODY study differs in its ability to estimate the adjusted odds of seroconversion associated with each disease, and each type of immunosuppression, which may influence immunotherapy decisions where there is equipoise in treatment benefit.

MELODY has also been able to assess the prevalence of psychological distress, defined as a combination of anxiety and depression measures, in immunosuppressed cohorts. The reported prevalence of psychological distress in the general population has varied during the different phases of the pandemic, with direct comparisons hampered by methodological differences in assessment^24^. From the MELODY participants who completed the PHQ-ADS, moderate to severe psychological distress was found in 7.4-16.9% of participants across the cohorts. However, the proportion of non-respondents was high, likely attenuating this estimate. As their antibody results were known to the participants when they completed their questionnaires, it is not known if, or how, this knowledge impacted on the psychological scoring. Interestingly, there are recognised mechanisms whereby psychological status may impact on adaptive immune responses, although this has not been studied in the setting of co-existing immunotherapy^25,26^. However, irrespective of causation, assessment of distress is important as observational data has shown an association between neuropsychiatric diagnoses and poorer outcome to COVID-19 infection^27^. Our data suggest there remains a significant prevalence of distress in the immunocompromised population which warrants recognition and consideration.

MELODY has recognisable limitations. First; participation required self-engagement in the community, and whilst the methodology offered broader and targeted reach to a vulnerable population with rare conditions, this does not necessarily overcome recognised barriers to research recruitment, such as the engagement and participation of people from ethnic minority backgrounds^28^. Although the number of participants is large enough to provide meaningful multi-ethnicity data, the relative predominance of white participants is noticeable across all cohorts. Second, is the use of a non-quantitative test that does not distinguish between no and very low SARS-CoV-2 IgG antibody levels, and that does not assess antigen-specific T-cell responses^29^. However, our aim was not to undertake a detailed immune response analysis, but rather to address whether mass antibody testing can discriminate the risk of severe COVID-19 in immunocompromised individuals. Further, whilst we have used one immunoassay, any approved assay testing for the presence of SARS-CoV-2 IgG could be used. We also acknowledge, we have not assessed all immunocompromised groups as defined by the WHO, e.g. persons with HIV, who will need further consideration^7^. Finally, although significant barriers have been overcome to deliver MELODY, data processing approvals have meant that a single intra-cohort comparison analysis was not possible, even though similar methodology and analysis has been applied to the different cohorts. Nevertheless, MELODY demonstrates a novel approach to recruiting and involving patients in rare disease research without using COVID-specific legislation, and its strengths include the large numbers of participants, and its ability to assess heterogeneity within broadly defined immunocompromised groups.

To conclude, the MELODY study demonstrates that immunocompromised individuals may be identified and reached via national disease registration services and the linkage of unique data sets. We plan to assess how the results of the home LFIA antibody tests relate to disease prevention by analysing SARS-CoV-2–related infection, hospitalisation and death rates in our immunocompromised cohorts. In this report, we corroborate risk factors associated with seronegativity in immunogenicity studies in distinct subcohorts of immunocompromised individuals, and describe commonality across cohorts. Given the influence of confounding characteristics, we show serological testing of immunosuppressed populations could provide personalised risk stratification not achieved by clinical characteristics alone. We also provide proof of principle of how this may be achieved in the community, and how serotesting this population may identify those individuals who may maximally benefit from pre-exposure prophylaxis^30^. Finally, and importantly, our data also supports the continued uptake of boosters in immunosuppressed patients, with seroconversion rates increasing with sequential vaccine doses. As a key challenge is to improve the vaccine-induced immunity of immunosuppressed individuals, it may also provide proof of principle for further bespoke booster schedules for this population, which could be guided by community antibody surveillance.

## Data Availability

The data used in this study are sensitive and will not be made publicly available.

## Author Contributions

All authors contributed to the design, execution and delivery of this cross specialty, collaborative study. Conceptualization (LL, SMc, FP, MW), Funding acquisition (GC,LL, PL, SL, SMc, FP, GPe, HT, HW, MW), Project administration (GPo), Methodology (MB, GC, LL, PL, SL, SMc, FP, GPe, HT, HW, MW), Data curation, cleaning and analysis (MB, JC, RH, SL, LM, FP, AT). Writing original draft (SL, GPe, FP, MW), Review and editing (MB, GC, JC, RH, LL, PL, SL, SMc, FP, GPe, AT, HT, HW, MW). All authors have seen and approved the final text and gave final approval of the version to be published.

## Declaration of interests

FP and PCL are recipients of an investigator-led grant from Vifor pharma. LL has received consulting fees from AstraZeneca, GSK, Pfizer, Biogen, Novartis and Alexion, which had no influence on the design, conduct or interpretation of this study. All other authors declare no competing interests.

## Data sharing

The data used in this study are sensitive and will not be made publicly available.

## Acknowledgements

This work uses data that has been provided by patients, the NHS and other health care organisations as part of routine patient care and support. The cancer and rare disease data are collated, maintained, and quality assured by the National Disease Registration Service, which are part of NHS Digital. The authors are grateful to all the transplant centres in the UK who contributed data on which the SOT analysis is based. Support for this work was also provided by Jeanette Aston, Peter Stilwell and Sean McPhail at the NDRS, and David Groves from the University of Nottingham. The authors would like to thank the patients who helped develop the study design. MW, SMc are supported by the National Institute for Health Research (NIHR) Biomedical Research Centre based at Imperial College Healthcare NHS Trust and Imperial College London. S.H.L. is supported by a Cancer Research UK Advanced Clinician Scientist Fellowship (A27179). HW is a National Institute for Health Research (NIHR) Senior Investigator and acknowledges support from NIHR Biomedical Research Centre of Imperial College NHS Trust, NIHR School of Public Health Research, and the NIHR Applied Research Collaborative North West London. Fiona Pearce is funded by the National Institute for Health Research (NIHR Advanced Fellow NIHR300863). The views expressed in this publication are those of the author(s) and not necessarily those of the NIHR, NHS or the UK Department of Health and Social Care.

The authors would like to extend their gratitude to the patient charities and their representatives involved in this work.

## Transparency Statement

MW affirms that the manuscript is an honest, accurate, and transparent account of the study being reported; and that no important aspects of the study have been omitted

## Notes

### Funding Statement

The study was funded by the Medical Research Council (MR/W029200/1), Kidney Research UK, Blood Cancer UK, Vasculitis UK and the Cystic Fibrosis Trust.

### Author Declarations

Ethical approval for MELODY (Mass Evaluation of Lateral flow immunOassays for the Detection of SARS-CoV-2 antibodY responses in immunosuppressed people) was granted by London Central Research Ethics Committee (21/HRA/4858) on 21 November 2021

