## Supplemental Data for "Antibody prevalence after 3 or more COVID-19 vaccine doses in 23,000 immunosuppressed individuals: a cross-sectional study from MELODY"

**Supplementary Data**

### **Additional Methods**

#### **NCRAS Rare Autoimmune Cohort selection for invitation to the MELODY study**

**Inclusion criteria:**

Patients aged ≥18 years on Nov 15, 2021 and resident in England who had a probable diagnosis of either anti-neutrophil cytoplasmic autoantibody (ANCA)-associated vasculitis or lupus or myositis or scleroderma or giant cell arteritis and were identified by the National Disease Registration Service (NDRS) using the following algorithms applied to Hospital Episode Statistics data. National data opt outs and NDRS opt outs were applied.

At least two ordinary or day case admissions in Hospital Episode Statistics (HES) between Apr 1, 2017 and Aug 31, 2021with diagnostic codes indicating one of the above diagnoses (see ICD-10 codes in table 1) AND EITHER

1. An admission between Apr 1, 2019 and Aug 31, 2021 indicating the administration of either rituximab or belimumab (OPCS code X921 or X951)

OR

2. At least one renal, respiratory or rheumatology outpatient clinic appointment (treatment function codes Rheumatology: 410, Respiratory: 340, Renal: 361) between Apr 1, 2019 and Aug 31, 2021.

Table 1 ICD-10 codes and description[1]

| **Condition** | **ICD-10 code** | **Description** |
| --- | --- | --- |
| AAV | M301 | Polyarteritis with lung involvement [Churg-Strauss] |
| AAV | M313 | Wegener granulomatosis |
| AAV | M317 | Microscopic polyangiitis |
| Lupus | M321 | Systemic lupus erythematosus with organ or system involvement |
| Lupus | M328 | Other forms of systemic lupus erythematosus |
| Lupus | M329 | Systemic lupus erythematosus, unspecified |
| Myositis | M332 | Polymyositis |
| Myositis | M609 | Myositis, unspecified |
| Myositis | M608 | Other myositis |
| Myositis | M331 | Other dermatomyositis |
| Myositis | M339 | Dermatopolymyositis, unspecified |
| Myositis | M360 | Dermato(poly)myositis in neoplastic disease |
| Scleroderma | M340 | Progressive systemic sclerosis |
| Scleroderma | M341 | CR(E)ST syndrome |
| Scleroderma | M348 | Other forms of systemic sclerosis |
| Scleroderma | M349 | Systemic sclerosis, unspecified |
| Giant Cell Arteritis | M315 | Giant cell arteritis with polymyalgia rheumatica |
| Giant Cell Arteritis | M316 | Other giant cell arteritis |

#### **NCRAS Blood Cancer Cohort selection for invitation to the MELODY study**

**Inclusion criteria:**

Patients aged ≥18 years and resident in England at the time of diagnosis who were registered in the National Cancer Registration Dataset [1] in 2019 or in the Rapid Cancer Registration Dataset [2] in 2020 or 2021 with a diagnosis of lymphoma or multiple myeloma (see definition below).

Table 1 ICD-O-3 codes and description [3]

| **ICD-O code** | **Description** |
| --- | --- |
| 9665/3 | Hodgkin lymphoma, nodular sclerosis, grade 1 |
| 9667/3 | Hodgkin lymphoma, nodular sclerosis, grade 2 |
| 9652/3 | Hodgkin lymphoma, mixed cellularity, NOS |
| 9650/3 | Hodgkin lymphoma, NOS |
| 9695/3 | Follicular lymphoma, grade 1 |
| 9691/3 | Follicular lymphoma, grade 2 |
| 9698/3 | Follicular lymphoma, grade 3 |
| 9690/3 | Follicular lymphoma, NOS |
| 9699/3 | Marginal zone B-cell lymphoma, NOS |
| 9671/3 | Malignant lymphoma, lymphoplasmacytic |
| 9761/3 | Waldenstrom macroglobulinaemia |
| 9673/3 | Mantle cell lymphoma |
| 9689/3 | Splenic marginal zone B-cell lymphoma (C42.2) |
| 9679/3 | Mediastinal large B-cell lymphoma (C38.3) |
| 9680/3 | Malignant lymphoma, large B-cell, diffuse, NOS |
| 9823/3 | B-cell chronic lymphocytic leukaemia/small lymphocytic lymphoma |
| 9732/3 | Multiple myeloma C42.1 |

**Data fields extracted**

- NHSNUMBER
- BIRTHDATEBEST (Date of birth)
- FORENAME
- SURNAME
- TUMOURID
- ICD03_REV2011 (ICD-O-3 morphology codes [3])

#### **Questionnaire response data processing**

The raw LM and RAD self-report data was pre-processed prior to the analysis. This involved three main cleaning steps: mapping free-text responses, grouping categories and applying hierarchy to treatment and diagnoses.

Multiple free-text fields were present in the questionnaire allowing respondents to describe ‘other’ vaccine types, treatments and diagnoses outside of the available options. Many of these included synonyms, descriptive text and misspellings of options that should have been specifically indicated. Additionally, many free-text entries contained immaterial information that allowed the response to be transferred from the other to ‘none’ group. These entries were mapped to the appropriate response following a free-text review by clinical specialists.

To reduce the total number of diagnoses and treatments levels for analysis and to incorporate those with low prevalence (N<50), a clinically informed grouping was applied. Following this, a hierarchy was used to assign those within multiple groups to a primary group for analysis (RAD: 5% within multiple diagnoses groups, 8% within multiple treatment groups. LM: 3% within multiple diagnoses groups, 3% multiple treatment groups). An exception was made here for multiple blood cancer diagnoses when the most recent diagnosis was chosen as the primary.

**Diagnosis grouping and multiple diagnoses hierarchy in rare autoimmune disease cohort.**

| **Group** | **Hierarchy rank** | **Condition** |
| --- | --- | --- |
| Small vessel vasculitis (SVV) | 1 | Primary CNS (Brain) vasculitis |
|  |  | Behcet's disease |
|  |  | Polyarteritis nodosa |
|  |  | Churg Strauss (or EGPA, eosinophilic granulomatosis with polyangiitis) |
|  |  | MPA (or microscopic polyangiitis) |
|  |  | GPA (or granulomatosis with polyangiitis / Wegener's) |
|  |  | ANCA vasculitis |
| Large vessel vasculitis (LVV) | 2 | Takayasu arteritis |
|  |  | Giant cell arteritis (temporal arteritis) |
| SLE | 3 | Systemic lupus erythematosus |
| Scleroderma | 4 | Scleroderma |
| Myositis | 5 | Polymyositis |
|  |  | Dermatomyositis |
| Other diagnoses | 6 | Other |
| None | 7 | None of these |

**Treatment grouping and multiple treatment hierarchy in rare autoimmune disease cohort.**

| **Group** | **Hierarchy rank** | **Treatment** |
| --- | --- | --- |
| Anti-CD20 (12 months) | 1 | Rituximab |
|  |  | Ofatumumab |
|  |  | Obinutuzumab |
|  |  | Ocreluzimab |
| Cyclophosphamide (12 months) | 2 | Cyclophosphamide |
|  |  | Alemtuzumab |
| Mycophenolate | 3 | MMF (Mycophenolate Mofetil, Mycophenolate Acid, Cellcept, Myfenax, Ceptava, Myfortic) |
| Azathioprine | 4 | Azathioprine (Imuran) |
| Methotrexate | 5 | Methotrexate Oral (Maxtrex, Jylamvo) |
|  |  | Methotrexate Injection/Infusion (Nordimet, Zlatal, Methofill, Metoject) |
| Other | 6 | Other |

**Diagnosis grouping and multiple diagnoses hierarchy in lymphoid malignancies cohort.**

| **Group** | **Hierarchy rank** | **Condition** |
| --- | --- | --- |
| Aggressive B-NHL | 1 (Most recent) | Diffuse Large B cell lymphoma |
|  |  | Burkitt lymphoma |
| Indolent B-NHL | 1 (Most recent) | Follicular lymphoma |
|  |  | Mantle cell lymphoma |
|  |  | Marginal zone lymphoma |
|  |  | Chronic lymphocytic lymphoma (CLL) |
| Plasma cell malignancies | 1 (Most recent) | Multiple myeloma |
|  |  | Plasmacytoma |
| Hodgkin Lymphoma | 1 (Most recent) | Hodgkin Lymphoma |
| Other diagnoses | 2 | Other lymphoma or leukaemia |
| None | 3 | None of these |

**Treatment grouping and multiple treatment hierarchy in lymphoid malignancies cohort.**

| **Group** | **Hierarchy rank** | **Treatment** |
| --- | --- | --- |
| Anti-CD20 (12 months) | 1 | Rituximab (Rituxan) |
|  |  | Obinutuzumab (Gazyva) |
| Ibrutinib / Acalabrutinib (3 months) | 2 | Ibrutinib (Imbruvica) |
|  |  | Acalabrutinib (Calquence) |
| Chemotherapy (3 months) | 3 | Chemotherapy |
|  |  | Methotrexate (Within 4 weeks) |
| Lenalidomide (3 months) | 4 | Lenalidomide (Revlimid) |
| Radiotherapy (3 months) | 5 | Radiotherapy |
| Autologous SCT (12 months) | 6 | Autologous stem cell transplant |
| Other Treatment | 7 | Other |

For the SOT group, a hierarchy was applied to the immunosuppression recorded with those receiving belatacept considered in the belatacept based group regardless of other immunosuppression drugs indicated. Those on antiproliferative or calcineurin were then considered by whether they had these in combination with each other. The ‘other’ group are those who selected a drug but did not select belatacept, antiproliferative, nor calcineurin. The ‘none’ group are those who selected ‘none of these’ on the survey.

### **Comparison of invited versus recruited participants**

To assess the sampled cohorts represent the whole population, a comparison between the baseline demographics of the analysed and invited cohorts were performed.

**a. Transplant cohort**

|  | **Analysis cohort** | **Invited cohort*** | **p-value** |
| --- | --- | --- | --- |
|  | **N (%)** | **N (%)** |  |
| **Total** | 9927 (100) | 47684 (100) |  |
| **Gender** |  |  |  |
| Male | 5426 (54.7) | 29064 (61.0) | <0.0001 |
| Female | 4494 (45.3) | 18592 (39.0) |  |
| Not reported | 7 | 28 |  |
| **Ethnicity** |  |  |  |
| White | 9268 (93.8) | 35876 (79.3) | <0.0001 |
| Asian | 351 (3.6) | 5817 (12.9) |  |
| Black | 134 (1.4) | 2639 (5.8) |  |
| Other | 133 (1.4) | 892 (2.0) |  |
| Not reported | 41 | 2460 |  |
| **Transplant type** |  |  | <0.0001 |
| Kidney only | 6591 (66.4) | 34229 (71.8) |  |
| Liver only | 1981 (20.0) | 8000 (16.8) |  |
| SPK/ Panc/ Islet/ SIK | 350 (3.5) | 2048 (4.3) |  |
| Heart only | 596 (6.0) | 1942 (4.1) |  |
| Lung (incl heart-lung) | 333 (3.4) | 1171 (2.5) |  |
| Other | 76 (0.8) | 294 (0.6) |  |
| **Graft number** |  |  |  |
| First graft | 8696 (87.6) | 41572 (87.2) | 0.26 |
| Re-graft | 1231 (12.4) | 6112 (12.8) |  |
| **Age (median, IQR)** | 60 (50-67) | 57 (46-66) | <0.0001 |

* People on the NHS BT register and ≥18 years and resident in England and Wales at the start of invitations

b. **Rare autoimmune disease group**

|  | **Analysis cohort** | **Invited cohort*** | **p-value** |
| --- | --- | --- | --- |
|  | **N (%)** | **N (%)** |  |
| **Total** | 6516 (100) | 32587 (100) |  |
| **Gender** |  |  |  |
| Male | 1447 (22.3) | 7267 (22.4) | 0.91 |
| Female | 5032 (77.7) | 25163 (77.6) |  |
| Not reported | 37 | 157 |  |
| **Ethnicity** |  |  |  |
| White | 6024 (93.1) | 26447 (81.7) | <0.0001 |
| Asian | 212 (3.3) | 2950 (9.1) |  |
| Black | 123 (1.9) | 2032 (6.3) |  |
| Other | 109 (1.7) | 945 (2.9) |  |
| Not reported | 48 | 213 |  |
| **Diagnosis** |  |  |  |
| Small vessel vasculitis | 1321 (20.3) | 4812 (14.8) | <0.0001 |
| Large vessel vasculitis | 1194 (18.3) | 8127 (24.9) |  |
| SLE | 2528 (38.8) | 13237 (40.6) |  |
| Scleroderma | 888 (13.6) | 3926 (12) |  |
| Myositis | 585 (9) | 2485 (7.6) |  |
| **Age (median, IQR)** | 65 (54.0 - 73.0) | 66 (52.0 - 77.0) | <0.0001 |

*Data extracted from hospital records with some discrepancies to self-report data. *Those eligible to be invited at the start of invitations (31/01/22).

c. **Lymphoid Malignancy Cohort**

|  | **Analysis cohort** | **Invited cohort*** | **p-value** |
| --- | --- | --- | --- |
|  | **N (%)** | **N (%)** |  |
| **Total** | 6593 (100) | 28748 (100) |  |
| **Gender** |  |  |  |
| Male | 3562 (54.8) | 16142 (56.8) | 0.0028 |
| Female | 2939 (45.2) | 12256 (43.2) |  |
| Not reported | 92 | 350 |  |
| **Ethnicity** |  |  |  |
| White | 5811 (95.2) | 23913 (89.0) | <0.0001 |
| Asian | 99 (1.6) | 1291 (4.8) |  |
| Black | 66 (1.1) | 760 (2.8) |  |
| Other | 130 (2.1) | 902 (3.4) |  |
| Not reported | 487 | 1882 |  |
| **Diagnosis** |  |  | <0.0001 |
| Aggressive B-NHL | 1364 (20.7) | 5929 (20.6) |  |
| Indolent B-NHL | 3401 (51.6) | 13655 (47.5) |  |
| Plasma cell malignancies | 1506 (22.8) | 7223 (25.1) |  |
| Hodgkin Lymphoma | 322 (4.9) | 1941 (6.8) |  |
| **Age (median, IQR)** | 69 (61.0 - 75.0) | 71 (60.0 - 78.0) | <0.0001 |

*Data extracted from hospital records with some discrepancies to self-report data. *Those eligible to be invited at the start of invitations (31/01/22).

### **Table S1. Antibody responses for all participants with 3 or more vaccines by IgM and IgG status**

| **Antibody response** | **Transplant cohort** | | **Rare autoimmune disease cohort** | | **Lymphoid malignancy cohort** | |
| --- | --- | --- | --- | --- | --- | --- |
|  | **N** | **%** | **N** | **%** | **N** | **%** |
| Negative | 1950 | 20 | 710 | 11 | 1142 | 17 |
| Negative – IgM only | 360 | 4 | 212 | 3 | 224 | 3 |
| Positive – IgG only | 5624 | 57 | 4191 | 64 | 3888 | 59 |
| Positive IgM + IgG | 1993 | 20 | 1403 | 22 | 1339 | 20 |
| **TOTAL** | **9927** | **100** | **6516** | **100** | **6593** | **100** |

### **Table S2. Logistic regression for antibody positivity^1^ in transplant recipients who had 3 or more vaccines**

|  | **N** | **Odds ratio (95% CI)** | **p-value** |
| --- | --- | --- | --- |
| **Age (10-year increase)** | 9233 | 0.70 (0.66-0.73) | <0.0001 |
| **Vaccines at test** |  |  | <0.0001 |
| 3 | 2472 | 1.00 |  |
| 4 | 5649 | 1.91 (1.70-2.15) |  |
| 5+ | 1112 | 2.76 (2.28-3.35) |  |
| **Immunosuppression** |  |  | <0.0001 |
| Antiproliferative and Calcineurin | 3080 | 1.00 |  |
| Antiproliferative only | 181 | 1.71 (1.11-2.64) |  |
| Calcineurin only | 1349 | 2.02 (1.66-2.45) |  |
| Other^2^ | 252 | 1.90 (1.31-2.75) |  |
| Antiproliferative and Calcineurin and Steroid | 2639 | 0.61 (0.53-0.70) |  |
| Antiproliferative and Steroid | 402 | 1.18 (0.90-1.54) |  |
| Calcineurin and Steroid | 1330 | 1.12 (0.94-1.32) |  |
| **Transplant type** |  |  | <0.0001 |
| Kidney only | 6165 | 1.00 |  |
| Liver only | 1814 | 1.27 (1.09-1.49) |  |
| SPK/ Panc/ Islet/ SIK | 323 | 0.69 (0.53-0.92) |  |
| Heart only | 545 | 0.94 (0.75-1.17) |  |
| Lung (including heart-lung) | 314 | 0.59 (0.46-0.77) |  |
| Other | 72 | 0.80 (0.46-1.38) |  |
| **Previous COVID-19 infection** |  |  | <0.0001 |
| No | 5283 | 1.00 |  |
| Yes | 3372 | 4.16 (3.65-4.75) |  |
| Unknown | 578 | 1.34 (1.09-1.66) |  |
| **Rejection** |  |  | 0.0002 |
| No | 9048 | 1.00 |  |
| Yes | 185 | 0.51 (0.36-0.71) |  |
| **Gender** |  |  | 0.026 |
| Male | 5103 | 1.00 |  |
| Female | 4130 | 0.89 (0.80-0.99) |  |
| **Vaccine type** |  |  | 0.0063 |
| mRNA + mRNA | 4077 | 1.00 |  |
| AZ + mRNA | 5156 | 0.86 (0.78-0.96) |  |
| **Graft number** |  |  | 0.022 |
| First graft | 8088 | 1.00 |  |
| Re-do | 1145 | 0.83 (0.71-0.97) |  |

^1^ Positive antibody result = IgG only or IgG + IgM

^2^ Other includes belatacept based and none. Those on other treatments and steroids are included in this group.

### **Table S3. Logistic regression for antibody positivity^1^ in participants with rare autoimmune rheumatic disease who had 3 or more vaccines**

|  | **N** | **Odds ratio (95% CI)** | **p-value** |
| --- | --- | --- | --- |
| **Age (10-year increase)** | 4866 | 0.90 (0.83 - 0.97) | 0.004 |
| **Vaccines at test** |  |  | <0.0001 |
| 3 | 1861 | 1.00 |  |
| 4 | 2710 | 1.62 (1.33 - 1.99) |  |
| 5+ | 295 | 3.28 (2.03 – 5.30) |  |
| **Immunosuppression** |  |  | <0.0001 |
| None^2^ | 1347 | 1.00 |  |
| Anti-CD20 (1 year) | 708 | 0.07 (0.05 – 0.10) |  |
| Cyclophosphamide (1 year) | 92 | 0.49 (0.24 – 0.98) |  |
| Mycophenolate (current) | 806 | 0.39 (0.28 – 0.55) |  |
| Azathioprine (current) | 447 | 0.68 (0.44 – 1.06) |  |
| Methotrexate (current) | 580 | 0.79 (0.53 – 1.18) |  |
| Other | 886 | 0.65 (0.46 – 0.91) |  |
| **Steroid for immunosuppression** | |  | <0.0001 |
| No | 2773 | 1.00 |  |
| Yes | 2093 | 0.66 (0.55 - 0.80) |  |
| **Diagnosis** |  |  | 0.0006 |
| Systemic lupus erythematosus | 2011 | 1.00 |  |
| Large vessel vasculitis | 520 | 1.31 (0.87 – 1.97) |  |
| Small vessel vasculitis | 1194 | 0.69 (0.55 - 0.87) |  |
| Scleroderma | 730 | 1.17 (0.84 - 1.61) |  |
| Myositis | 396 | 1.18 (0.84 - 1.67) |  |
| Other diagnosis | 15 | 0.51 (0.13 – 2.04) |  |
| **Previous COVID-19 infection** |  |  | <0.0001 |
| No | 2747 | 1.00 |  |
| Yes | 1756 | 2.32 (1.87 - 2.87) |  |
| Unknown | 363 | 1.59 (1.10 - 2.30) |  |

^1^ positive antibody result = IgG only or IgG + IgM

^2^ includes hydroxychloroquine

### **Table S4. Logistic regression for antibody positivity^1^ in participants with lymphoid malignancies who had 3 or more vaccines**

|  | **N** | **Odds ratio (95% CI)** | **p-value** |
| --- | --- | --- | --- |
| **Age (10 year increase)** | 5737 | 0.76 (0.71 - 0.82) | <0.0001 |
| **Vaccines at test** |  |  | <0.0001 |
| 3 | 1073 | 1 |  |
| 4 | 4101 | 1.37 (1.14 - 1.66) |  |
| 5+ | 563 | 1.96 (1.42 - 2.71) |  |
| **Immunosuppression** |  |  | <0.0001 |
| None | 3071 | 1 |  |
| Chemotherapy (3 months) ^2^ | 482 | 0.12 (0.09 - 0.16) |  |
| Anti-CD20 | 370 | 0.05 (0.04 - 0.06) |  |
| Radiotherapy (3 months) | 50 | 0.21 (0.12 - 0.39) |  |
| BTKi (3 months) | 180 | 0.16 (0.12 - 0.23) |  |
| Thalidomide analogue (3 months) | 358 | 0.34 (0.21 - 0.57) |  |
| ASCT (12 months) | 94 | 0.29 (0.14 - 0.58) |  |
| Other | 1132 | 0.19 (0.16 - 0.23) |  |
| **Diagnosis** |  |  | <0.0001 |
| Hodgkin Lymphoma | 463 | 1 |  |
| Indolent B-NHL | 2539 | 0.58 (0.43 - 0.77) |  |
| Aggressive B-NHL | 944 | 0.50 (0.37 - 0.69) |  |
| Plasma cell malignancies | 1194 | 3.19 (2.20 - 4.62) |  |
| Other NHL | 524 | 0.67 (0.47 - 0.96) |  |
| Other diagnosis | 73 | 0.84 (0.41 - 1.71) |  |
| **Previous COVID-19 infection** |  |  | <0.0001 |
| No | 3548 | 1 |  |
| Yes | 1769 | 1.93 (1.63 - 2.29) |  |
| Unknown | 420 | 1.53 (1.13 - 2.08) |  |

^1^ positive antibody result = IgG only or IgG + IgM

### **Table S5. Mental health comorbidities by antibody positivity in the transplant cohort**

|  | **Antibody negative**  **N (%)** | **Antibody positive**  **N (%)** | **p-value** |
| --- | --- | --- | --- |
| Total | 2310 (23.3%) | 7617 (76.7%) |  |
| Depression |  |  |  |
| No | 2107 (23.0%) | 7049 (77.0%) | 0.041 |
| Yes | 203 (26.3%) | 568 (73.7%) |  |
| Anxiety |  |  |  |
| No | 2100 (23.3%) | 6918 (76.7%) | 0.93 |
| Yes | 210 (23.1%) | 699 (76.9%) |  |
| Psychiatric disorder |  |  |  |
| No | 2296 (23.3%) | 7577 (76.7%) | 0.76 |
| Yes | 14 (25.9%) | 40 (74.1%) |  |
| Any mental health comorbidity |  |  |  |
| No | 2011 (23.1%) | 6681 (76.9%) | 0.42 |
| Yes | 299 (24.2%) | 936 (75.8%) |  |

### **Table S6. Mental health comorbidities by antibody positivity in the rare disease cohort**

|  | **Antibody negative**  **N (%)** | **Antibody positive**  **N % (%)** | **p-value** |
| --- | --- | --- | --- |
| Total | 922 (14.2%) | 5594 (85.8%) |  |
| Depression |  |  |  |
| No | 827 (14.5%) | 4892 (85.4%) | 0.061 |
| Yes | 95 (12.0%) | 702 (88.0%) |  |
| Anxiety |  |  |  |
| No | 832 (14.5%) | 4894 (85.5%) | 0.020 |
| Yes | 90 (11%) | 700 (89%) |  |
| Psychiatric disorder |  |  |  |
| No | 919 (14.2%) | 5554 (85.8%) | 0.257 |
| Yes | 3 (7.0%) | 40 (93.0%) |  |
| Any mental health comorbidity |  |  |  |
| No | 791 (14.7%) | 4588 (85.3%) | 0.006 |
| Yes | 131 (11.5%) | 1006 (88.5%) |  |

### **Table S7. Mental health comorbidities by antibody positivity in the Lymphoid Malignancy Cohort**

|  | **Antibody negative** | **Antibody positive** | **p-value** |
| --- | --- | --- | --- |
|  | **N (%)** | **N (%)** |  |
| Total | 1366 (20.7%) | 5227 (79.3%) |  |
| Depression |  |  |  |
| No | 1306 (20.8%) | 4979 (79.2%) | 0.633 |
| Yes | 60 (19.5%) | 248 (80.5%) |  |
| Anxiety |  |  |  |
| No | 1296 (20.9%) | 4915 (79.1%) | 0.261 |
| Yes | 70 (18.3%) | 312 (81.7%) |  |
| Psychiatric disorder |  |  |  |
| No | 1364 (20.8%) | 5207 (79.2%) | 0.278 |
| Yes | 2 (9.1%) | 20 (90.9%) |  |
| Any mental health comorbidity |  |  |  |
| No | 1261 (20.9%) | 4776 (79.1%) | 0.289 |
| Yes | 105 (18.9%) | 451 (81.1%) |  |

### **Table S8. Logistic regression results for impact of psychological distress on antibody positivity^1^ by disease cohort**

| **Cohort** | **Psychological**  **distress** | **N** | **Odds ratio (95% CI)** | **p-value** |
| --- | --- | --- | --- | --- |
| **Transplant** | No | 6715 | 1 | <0.0001 |
|  | Yes | 951 | 0.64 (0.54-0.76) |  |
|  | Not reported | 1567 | 0.86 (0.74-0.99) |  |
| **Rare autoimmune disease** | No | 3270 | 1 | 0.0025 |
|  | Yes | 664 | 1.17 (0.88 -1.55) |  |
|  | Not reported | 932 | 0.70 (0.56-0.88) |  |
| **Lymphoid malignancy** | No | 4466 | 1 | 0.39 |
|  | Yes | 356 | 0.87 (0.64 -1.17) |  |
|  | Not reported | 915 | 0.89 (0.73 -1.08) |  |

^1^ positive antibody result = IgG only or IgG + IgM
